# Assessing the Tendency of 2019-nCoV (COVID-19) Outbreak in China

**DOI:** 10.1101/2020.02.09.20021444

**Authors:** Qinghe Liu, Zhicheng Liu, Deqiang Li, Zefei Gao, Junkai Zhu, Junyan Yang, Qiao Wang

**Author notes:** Correspondence to Q. Wang.

## Abstract

Since December 8, 2019, the spread of COVID-19 is increasing every day. It is particularly important to predict the trend of the epidemic for the timely adjustment of the economy and industries. We performed the trends of 2019-nCoV (COVID-19) in China based on Flow-SEHIR model.

The results show that the basic reproductive numbers ***R***_***0***_ of COVID-19 is 3.56 (95% CI: 2.31 – 4.81). The number of daily confirmed new cases reaches the inflection point on Feb. 6 – 10 outside Hubei. For the maximum of infected cases number, the predicted peak value in China except Hubei was estimated to be 13806 (95% CI: 11926 - 15845). The peak arrival time is on March 3 - 9. The peak cumulative number of patients in Hubei was estimated to be 63800 (95% CI: 59300 - 76500). The temporal number of patients in most areas of China outside Hubei will peak from March 12 to March 15. The peak values of more than 73.5% provinces or regions in China will be controlled within 1000.

## 1. INTRODUCTION

Coronavirus [1] is a single stranded RNA virus, which is spherical in shape and has petal-like spines. It usually causes mild respiratory infections and can sometimes be fatal. Since its first discovery and identification in 1965, there have been three large-scale outbreaks of highly treatable infectious diseases caused by coronavirus, namely, SARS in mainland China in 2003, MERS in Saudi Arabia in 2012 and MERS in South Korea in 2015 [2].

At the beginning of December, the first patient infected by a new type of coronavirus was found in Wuhan, Hubei, which is a triffic center located in the central part of China. Then similar symptoms were found in the same city. After being infected, the main symptoms of patients in the early stage include low fever and fatigue, which were similar to the symptoms caused by general influenza, more seriously, some infected patients are asymptomatic, so it spreads in a large area without people’s awareness. On January 12, 2020, the World Health Organization officially named the new coronavirus causing pneumonia in Wuhan as “ 2019 new coronavirus [3] (SARS-CoV-19)”. On January 22, 2020, researchers speculated that the new coronavirus might come from wild bats [4]. During the Spring Festival of 2018 (from February 1 to March 12, 2018), the total estimated number of passenger trips in mainland China is about 2.97 billion [5]. As of 9am on February 7, 2020, the cumulative reports of 31 provinces (districts and municipalities) received by the National Health Commission of the People’s Republic of China [6] are shown in Table 1. In the table, Hubei Province’s data of confirmed cases is temporarily excluded since Hubei Province, as a severe epidemic area, has the possibility of a large backlog of confirmed cases, combined with the shortage of medical resources and the uncertainty of the diagnosis method, which makes the data in Hubei more unreliable than other provinces. Among these reports, 22112 cases of pneumonia with new coronavirus infection were found in Hubei Province [7].

**Table 1.** Real time epidemic situation in China by 9am February 7. (*Data source: data released by the National Health Commission of the People’s Republic of China and the Health Commissions of all provinces and autonomous regions*)

| Cumulative total | Confirmed cases<br>(except Hubei) | Death<br>cases | Cured<br>cases | Suspected<br>cases |
| --- | --- | --- | --- | --- |
| 31 provinces of China | 9049 | 637 | 1543 | 27657 |

The serious epidemic has brought many urgent problems. When will the spread of the epidemic turn to stabilize? When the growth of the number of confirmed cases can stop? To answer these questions, we perform an empirical analysis of the epidemic trend. In section 2, the most up-to-date literature by 7 Feb 2020 related to COVID-19 epidemic situation will be discussed and briefly analyzed. Section 3 presents Flow-SEHIR model in details. Section 4 gives the simulations and analysis. Finally, section 5 is the conclusions.

## 2. RELATED ISSUES

On January 27, 2020, B. Tang *et al* [8] established a transmission dynamics model based on the transmission mechanism of new coronavirus in Wuhan, isolation and other strategies. They thought the reproduction number of infection is 3.8 and the intervention measures can reduce the reproduction number of infection and the risk of transmission. Based on the epidemic data from Jan. 10 to Jan. 22, they predicted the basic reproduction number of infection of novel coronavirus in Wuhan is 6.47 (95% CI: 5.71 – 7.23).

On January 27, 2020, J. A. Backer *et al* [9] estimated that the average incubation period should be 5.8 days (95% CI: 4.6-7.9 days) and the range of variation be 1.3-11.3 days, according to the travel records and symptoms of 34 confirmed cases found outside Wuhan. From which they considered that 14 days of isolation will ensure that no symptoms will occur. In addition, they compared the COVID-19 with SARS and MERS.

On January 31, 2020, J. Wu *et al* [10] used data from Dec 31, 2019, to Jan 28, 2020 and thought that the basic reproduction number of infection was estimated by using Markov Chain Monte Carlo methods and presented by the the resulting posterior mean and 95% CI. They estimated that the basic reproduction number of the infected in Wuhan in 2019 was 2.68 (95% CI: 2.47-2.86). In addition, as of January 25, the number of infections was 75815 (95% CI: 37304-130330). The epidemic doubling time was 6.4 days (95% CI: 5.8-7.1).

On February 2, 2020, Adam J Kucharski *et al* [11] from London School of Hygiene & Tropical Medicine used a stochastic transmission model, based on confirmed cases in Wuhan and exported cases originating in Wuhan and estimated that the reproduction number of infection from mid-December to mid-January 2020 fluctuated between 1.6-2.9.

Epidemiological study is often an afterwards research, e.g. [12]. However, there are many uncertainties and risks in the development of COVID-19, so we need to provide some suggestions from the data analysis work in time.

On Feb.9, 2020, Li *et al* [13]simulated the dynamics of the COVID-19 and their results show that after the the traffic blockage in Jan.23,2020, the reproductive number in Wuhan will be 2.5, lower than before. The confirmed cases increased in linear not in exponential, which can confirm the effectiveness of strict control measures.

On Feb.12, 2020, Lin *et al* [14] provide estimates of the daily trend in the size of the epidemic in Wuhan and found the number of cases that should be reported in Wuhan by January 11, 2020 is 4090 (95% CI: 3975 – 4206) and 56833 (95% CI: 55242 – 58449) by February 9, 2020.

Since the outbreak of the epidemic, many scholars have concentrated in Wuhan and other areas, and studied the impact of basic reproduction number of infection and traffic interruption. However, the outbreak of the epidemic has a huge impact on the economy and people’s livelihood. Predicting the peak arrival time and evaluating the effectiveness of the control measures will play a decisive role in ensuring the smooth operation of the social economy. In this paper, two main areas (Hubei and other regions outside Hubei in China) are focused on for larger scale analysis. Li *et al* [15] conducted an in-depth analysis of the effects of traffic blockage and medical quarantine. It is shown that the quarantine is very effective for epidemic control, which can reduce the number of infections by nearly 90%.

## 3. COVID-19 TRANSMISSION MODEL: FLOW-SEHIR

### 3.1. SEIR Model

The SEIR model is introduced in 1984 by Aron and Schwartz [16], where S, E, I and R referring to the Susceptibles, Exposed, Infective, and Recovered, respectively. The followed SEIR model added the Exposed [17][18]. In the epidemic transmission, when susceptible ones are infected, most of them will neither immediately get sick nor have symptoms, but patients in the exposed period are also infectious, and during the recovery they will possibly be immune. Finally, the incubation period and the time-varying propagation rate are respectively investigated in [19].

### 3.2. Flow-SEHIR Model

Two main assumptions are adopted in this model:

1. The interaction of people flow between domestic and foreign areas is ignored.
2. During the transmission of virus, the probability of infection in contact with each person is equal.

Considering the special communication feature in Chinese Spring Festival and the effectiveness of nucleic acid testing is questionable, we add the factor of flow to reflect the mobility of the population and use *H* denoting ‘Hidden’, which means patients not detected by nucleic acid test. This forms our Flow-SEHIR model:

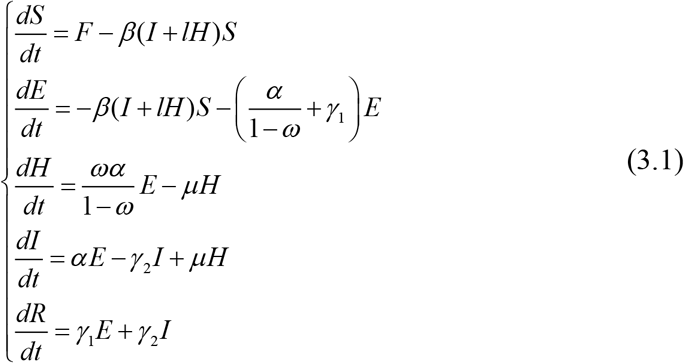

Here the flow *F*(t) rewrites the standard SEIR model by the first equation of (3.1). The explanation of all these parameters in this model is listed in Table 2.

**Table 2.** List of Symbols

|  |  |
| --- | --- |
| $S$ | Number of susceptible people |
| $E$ | Number of exposed people |
| $H$ | Number of false-negative people |
| $I$ | Number of infective people |
| $R$ | Number of recovered people |
| $\beta$ | Rate of those susceptible and being infected. |
| $\alpha$ | Rate of those exposed and being infectious. |
| $\gamma_1$ | Transmission rate to the recovered from the exposed. |
| $\gamma_2$ | Transmission rate to the recovered from the infectious. |
| $\gamma_3$ | Transmission rate to the recovered from the false-negative. |
| $\mu$ | Daily detection rate of false-negative patients. |
| $\omega$ | The proportion of false-negative in the actual population. |
| $l$ | The ratio of transmission of the false-negative to that of the confirmed. |

### 3.3. Flow and the Network of Cities

The outbreak of COVID-19 occurred around the Spring Festival in China, involving nearly a billion people moving around the country. Based on this feature, we constructed a (time-varying) directed graph to characterize the traffic flows across the nation, in which each node stands for a province (or city). The flow intensity *F*_*ji*_from *i*-th node to *j*-th one indicates the strength of the population flow. To perform this model, we collected population migration data from one of the biggest online location based service providers, Baidu, which provides daily intra-province population migration strength. In *i*-th city or province, this floating population is defined as follows:

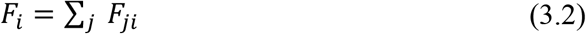

in which *F*_*ji*_= (1− *τ*)*δ*_*ji*_*S*_*i*_ characterizes the flow components from *j*-th node to *i*-th node, where the natural flow factor *δ ji* is independent of control policy of traffic blockage, and the traffic blockage factor *τ* is introduced to measure the control factor. Both the factors are between 0 and 1. For the latter, the higher the degree of traffic blockage over traffic flow implemented, the greater the flow of restriction follows. When *τ*=0, the traffic flow is in the uncontrolled state.

### 3.4. Practical Consideration

In our Flow-SEHIR model, the transmission process of the COVID-19 can be divided into the following four main stages:

1. The outbreak is potentially spreading, but was not been aware of;
2. Starting from the first case was officially reported since December 8, 2019;
3. Offcial confirmation (including transmission control in key cities, such as Wuhan, Hubei);
4. The epidemic is under control and most of patients have been recovered. Return trend, school opens days are mostly concentrated in this stage.

Our model will simulate stage 2, 3 and 4. Assume that in each stage, the recovery rates are relatively consistent, in terms of the natural recovery of self-recovered and medical treatment. But at each stage, the infection rate and the population can vary dramatically. For the first two stages, the spread of the virus satisfies the exponential distribution. For the third stage, it is found that there is a large number of population flows in and out because of the transportation during Spring Festival. For the third stage, there are a large number of population flows out from Wuhan, because of the Spring Festival as well as the panic. In this case, *F* will increase sharply. After then, traffic blockage will make *F* decrease. Due to self-protection and quarantine, the quarantine factor *ε* increases, then *β* decline. For the recovery rate parameters, we collected the empirical data related to pneumonia as the prior distributions.

From the second generator approach [20], we obtain the following expression for the basic reproductive number:

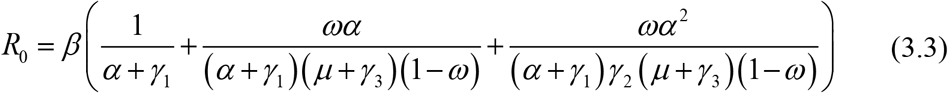

### 3.5. Data Source and Processing

In this study, we use official open dataset from CDC of the United States, World Health Organization, National Health Commission of China, and Baidu migration API. The details of data source are shown in Appendix 1.

#### National Health Commission of the people’s Republic of China

The national health commission of the People’s Republic of China regularly releases information on the epidemic situation in different parts of the country, which is officially released as public data, including local real-time diagnosis data, suspected data, cure data, etc.

#### Centers for Disease Control and Prevention (*CDC*)

CDC of the United States provides daily updated data on diagnosis, death and cure in various regions of China.

#### World Health Organization (*WHO*)

WHO provides timely official pneumonia of COVID-19, such as official naming, epidemic analysis, etc.

#### Baidu migration API

Baidu migration data is free and open traffic flow data obtained on baidu platform, including traffic intensity, migration ratio, etc.

We stress here that on February 12, 2020, the number of newly diagnosed patients in Hubei increased by about 14000, because that the new diagnosis specification is issued. For expanding the treatment scale, the confirmation of the disease will not only depend on the nucleic acid test, which will help to greatly reduce the proportion of false negative patients. Considering that this is a policy mutation, it is assumed that only this day is abnormal in terms of increment, and the normal increment level is restored after February 12.

## 4. MAIN RESULTS OF FLOW-SEHIR MODEL

### 4.1. Estimate of *R*_*0*_

The basic reproduction number, ***R***_***0***_, is an important index of epidemiology. It refers to the average value of how many people an infected person can transmit the virus to through natural transmission without external intervention.

Before January 20, 2020, when human-to-human transmission of COVID-19 was officially confirmed, people had not realized the infection of the epidemic and the SARS-CoV-2 was in a state of natural transmission. By estimating the trend in the early stages of the epidemic, combining equation (3.3), the basic reproductive numbers ***R***_***0***_ of COVID-19 is 3.56 (95% CI: 2.31 – 4.81).

### 4.2. Estimate of the Peak of Confirmed Cases

We define the peak value as the peak of *I*(t), which is the cumulative number of infected cases. After this peak point, the temporary number of cases gradually decrease and the epidemic subsides. We first simulated the epidemic trend of 34 provinces of China, and predicted the peak number of confirmed cases and its arrival time in each area, as shown in figure 2 and 3. On average, the peak number of real-time confirmed cases of Hubei is predicted to reach 63800, and its arrival time is on March 3, 2020. Tibet has both the minimum peak value and the earliest peak arrival time. According to the data up to Feburary 12, 2020, the number of confirmed cases in Tibet is only 1, and there is no suspected patient. On the one hand, it is shown that the transmission speed is very high, on the other hand, the rapid development of traffic caused the rapid development of cross transmission. The predicted peak values of different areas are divided to different intervals, which is listed in table 3. Except for Hubei, there are no other provinces that the peak value is more than 2000.

**Table 3.** Predicted peak number of confirmed cases of each province except Hubei with *τ* = 0.9, *ε* = 0.25 and the mean values of recovery rate and infection rate.

| Provinces (autonomous regions, or special administrative regions) | Peak value interval |
| --- | --- |
| Guangdong, Henan, Zhejiang, Anhui, Hunan | [1000, 2000] |
| Jiangxi, Jiangsu, Shandong, Chongqing, Heilongjiang, Sichuan | [500, 1000] |
| Beijing, Hebei, Shanghai, Fujian, Guangxi, Shaanxi, Guizhou | [200, 500] |
| Hainan, Yunnan, Tianjin, Shanxi, Liaoning, Jilin, Gansu, Xinjiang, Inner Mongolia, Ningxia, Hongkong, Taiwan, Qinghai, Macau, Tibet | <200 |

**Figure 1.**
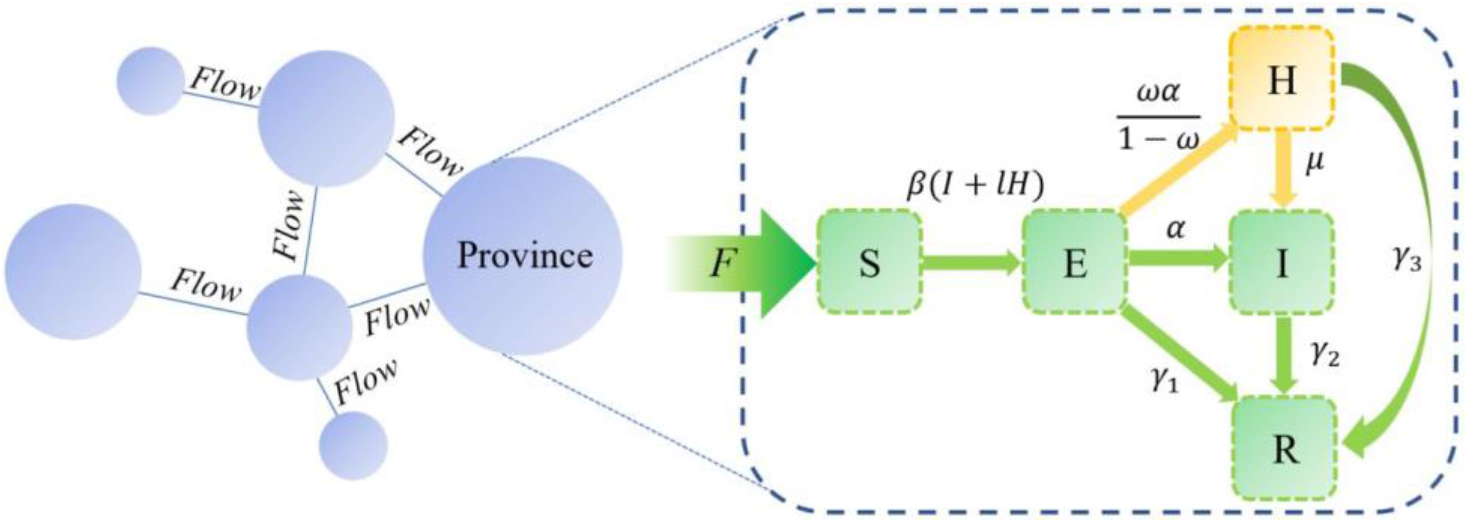
Flow-SEHIR Model Diagram.

**Figure 2.**
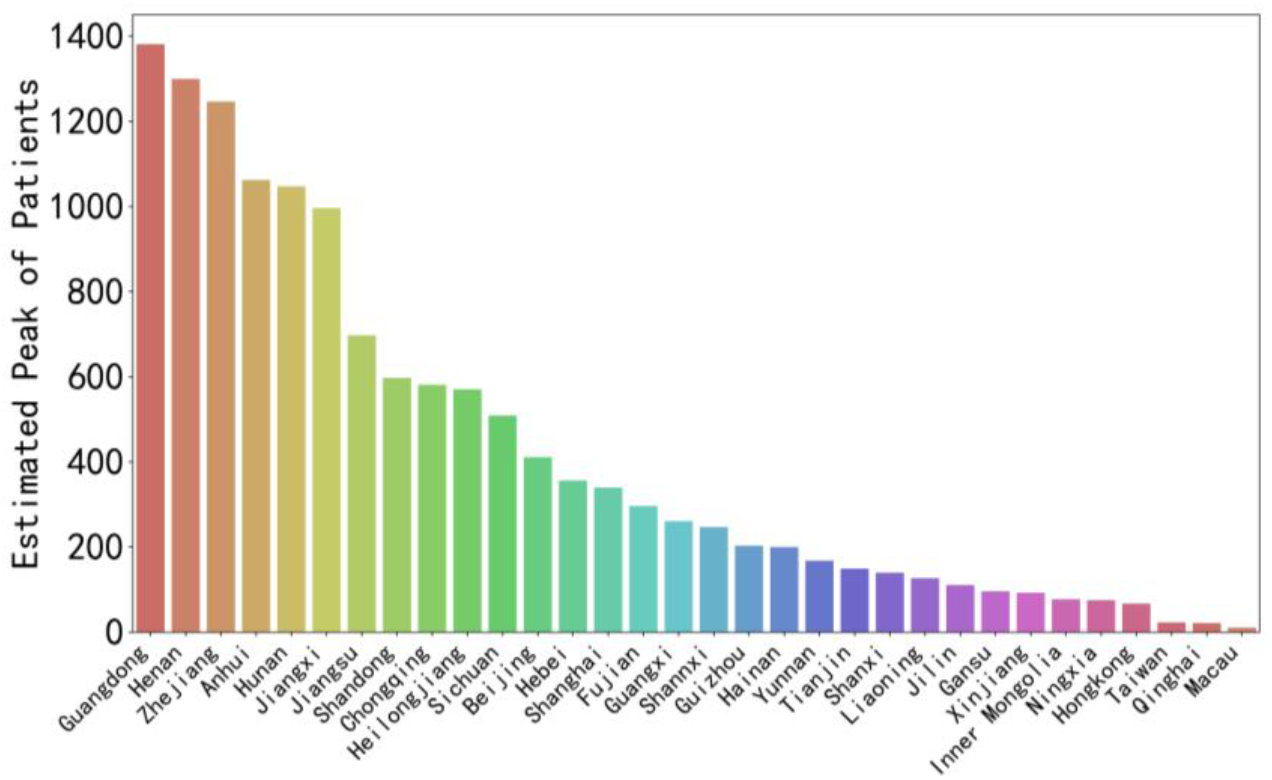
Predicted peak number of patients in each province of China, the x-axis corresponds to different geographical areas and the y-axis is the logarithm calculation of the predicted number of people.

**Figure 3.**
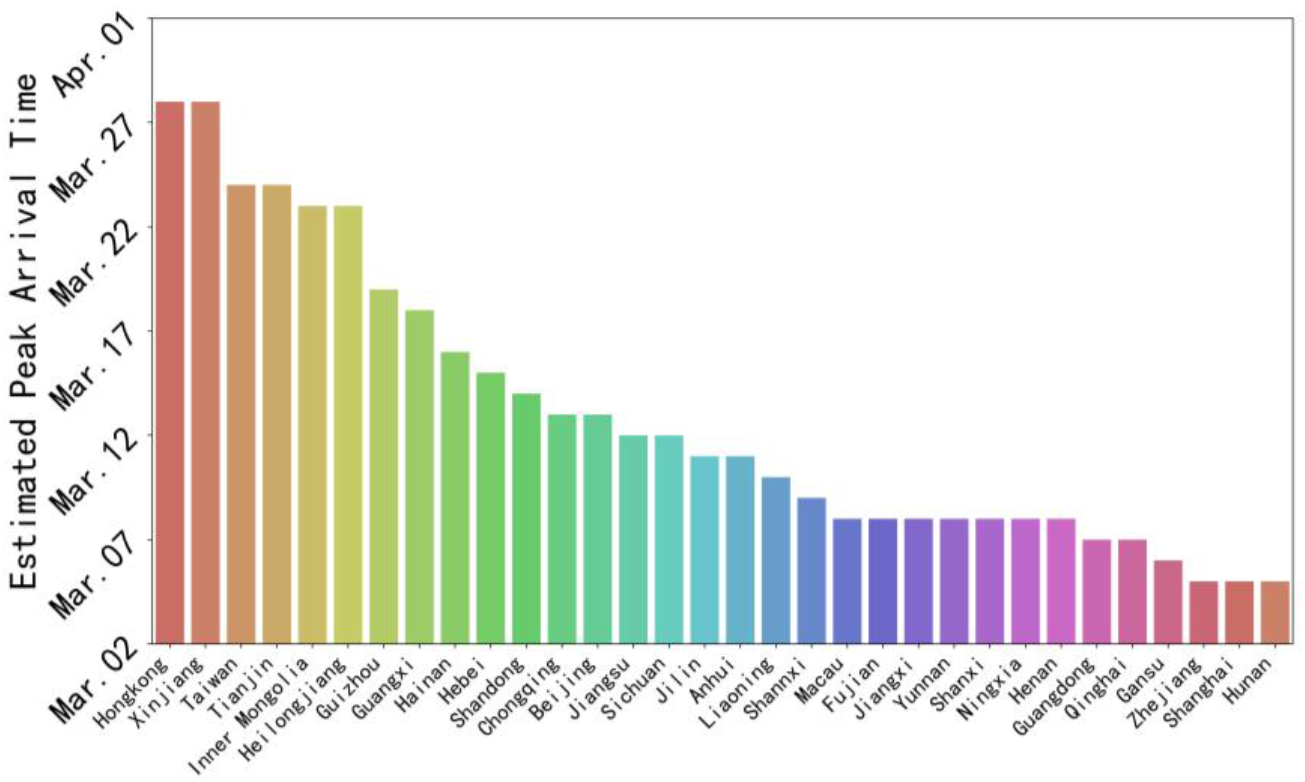
Predicted arrival time of peak number of confirmed cases in each province of China, the x-axis corresponds to different geographical areas and the y-axis is the prediction of peak arrival time at average level.

Table 3 distinguishs Hubei Province as a single category to reflect the corresponding relationships and differences. In terms of peak values, Hubei is far higher than other regions. Zhejiang, Guangdong, Henan, Hunan and Chongqing are the places most likely to become the next worst-hit areas, and the relevant departments should pay much attention to them. The peak value of more than 73.5% provinces or regions of China is predicted to be stable within 1000.

It is estimated that the peak arrival time of Hubei will be at the begining of March, which is earlier than other provinces in China. Therefore, it is very urgent to analyze the progress of the disease in Hubei. Then, comparative analysis and discussion on more details of Hubei are conducted.

### 4.3. Simulation of Confirmed Cases

According to the results, the potential population affected by the epidemic will be stable in March 2020. The epidemic situation in Hubei will not be completely controlled and eliminated until June 2020. Except that the trend of except-Hubei is basically consistent with that of Hubei, the change of the epidemic situation of except-Hubei is higher than that in Hubei. The trend of the confirmed cases’ number in Hubei is steeper, which is consistent with the characteristics of a diffusion center, the change of which time is earlier and drastic. While comparing with the center of epidemics, the periphery presents the characteristics of postposition and slowness. The trend is usually steeper and ahead of time in the center of the epidemic compared with other regions. On the one hand, it is related to the timely protection measures that have taken to other areas. On the other hand, it is due to the information lag of the central location. Similarly, according to the data, compared with other regions in Hubei Province, Wuhan also shows obvious prepositiveness and steepness. In addition, the return trend will not cause the next peak of illness, and is relatively safe, due to the improvement of people’s awareness of isolation and protection, as well as the importance of epidemic management in various industries.

The errors between the predicted data and the real data in Beijing, Shanghai, Fujian and Guangdong are shown in figure 4. Here we difine the inflection point as the peak value of new infections number per day, and after the inflection point, the number of infected people will grow slowly and the epidemic will be gradually controlled. We can get the inflection point after first-order difference calculation of the cumulative cases trend, which is expected to be on February 6 - February 10.

**Figure 4.**
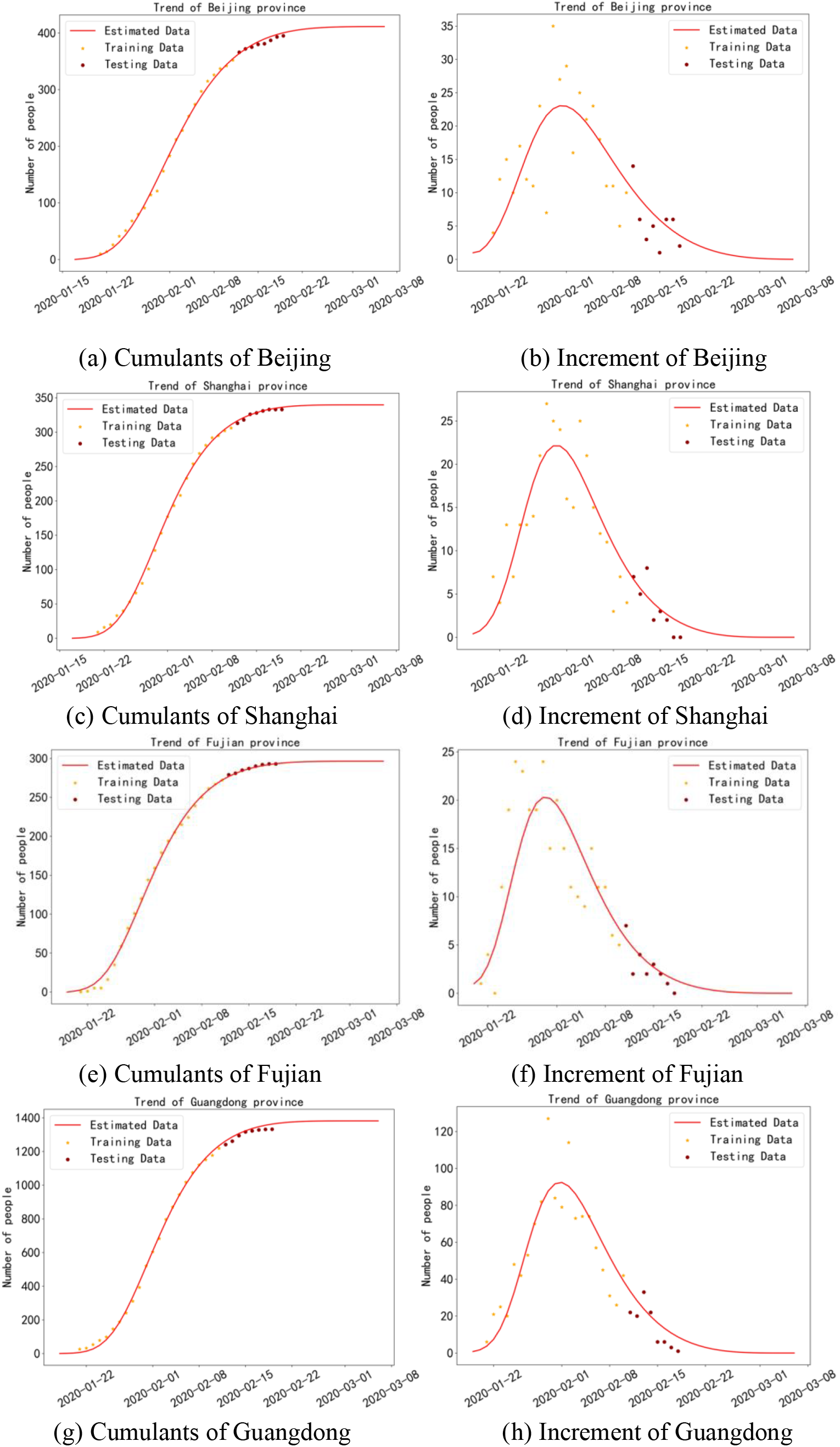
Comparisons between the estimated infective data and the real infective data, the x-axis is date and the y-axis is the number of infective people.

**Figure 6.**
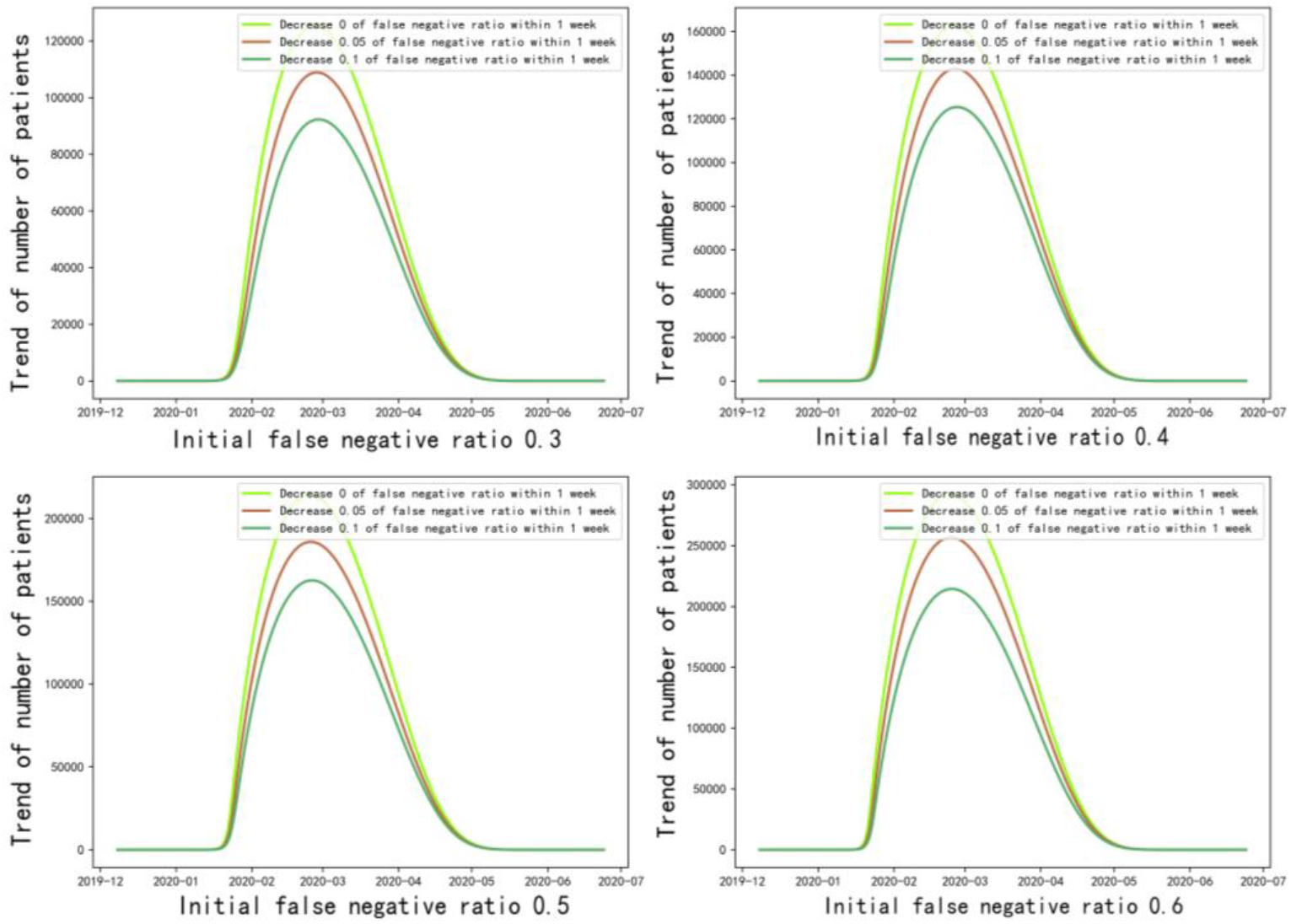
Estimation of the real number of patients with different decrease on false negative ratio (0%, 5%, 10%) within a week.

We took the upper quantile of 0.9 as the cure rate, and took the upper quantile of 0.1 as the prevalence rate, to represent a better case for simulation; and we took the upper quantile of 0.1 as the cure rate, took the upper quantile of 95% as the prevalence rate, to represent a worse case for simulation, and the results are shown in Table 4. The peak time fluctuations of Hubei and other regions are respectively about Feburary 3, 2020 (95% CI: Feburary 27 – March 18) and March 13, 2020 (95% CI: March 12 – March 15).

**Table 4.** Simulation results of epidemic situation in China except Hubei

|  | Peak value time | Peak value |
| --- | --- | --- |
| Worse case | 2020. 03.15 | 15845 |
| Average case | 2020. 03.13 | 13806 |
| Better case | 2020. 03.12 | 11926 |

### 4.4. Estimate of the real patients number in Hubei province

In order to study the effect of false negative proportion on the scale of the peak of confirmed cases, we simulated different false negative proportion, and discussed the final effect of the decrease of false negative proportion on the scale of the epidemic at different time points.

In figure 5, the results show that if the false negative rate decreased by 5% in one week, the peak number of patients would slow down by 11.62%. If the proportion decreased by 10% in one week, the peak number of patients would decrease by 21.91%. It can be seen that timely detection of false negative population is more effective for epidemic control.

## 5. CONCLUSION

We performed simulation analysis of COVID-19 based on Flow-SEHIR model.

From analysis, we draw the following conclusions:

1. We used the experimental results before January 20, 2020 (before the official confirmation that COVID-19 has the phenomenon of human-to-human transmission) to simulate the natural transmission of the SARS-CoV-2. The basic reproductive numbers ***R***_***0***_ was estimated to be 3.56 (95% CI: 2.31 – 4.81).
2. The peak of areas except Hubei in China will come at March 13, 2020 (95% CI: March 12 –March 15). The peak values of more than 73.5% of provinces and regions in China will be controlled within 1000. Rather than larger outbreak, the estimated number of infective patinets in Hubei(in which Wuhan is situated) presents a front and steep feature, manifesting the overall trend that the epidemic tends to be consistent.
3. The predicted peak value in China except Hubei is 13806 (95% CI: 11926 - 15845). The estimated peak value in Hubei is estimated to be 63800 (95% CI: 59300 - 76500), and its arrival time is on March 3, 2020 (95% CI: Feburary 27 – March 18). The predicted peak value of China is 85500 (95% CI: 76700 - 97500). It will take about 1.5-2 months from the peak to the end of the epidemic. Due to the people’s sense of isolation and safety protection, the return trend will not have a lot of impact on the epidemic, which will continue to show a downward trend after February 10.
4. Suppose that the transmission rate of the false negative patient is twice that of the confirmed patient, if the false negative rate decreased by 5% in one week, the peak number of patients will slowed down by 11.62%. If the proportion decreased by 10% in one week, the peak number of patients will decreased by 21.91%, which shows timely detection of false negative population is more effective for epidemic control.

## Data Availability

All the data are publicly available from the network

http://qianxi.baidu.com/

http://www.nhc.gov.cn

http://ynswsjkw.yn.gov.cn/wjwWebsite/web/index

http://wjw.beijing.gov.cn/

http://wjw.hubei.gov.cn/

## Abbreviations

NCP: Novel Coronavirus Pneumonia
COVID-19: Coronavirus Disease 2019
SARS-CoV-2: Severe Acute Respiratory Syndrome Coronavirus 2
CI: Confidence Interval
SEIR: Susceptible-Exposed-Infectious-Recovered

## Declarations

### Ethics approval and consent to participate

Not applicable.

### Consent for publication

Not applicable.

### Availability of data

The datasets used and analyzed during the current study is available from open resources.

### Authors’ Contributions

Conceived and designed the experiments: Qinghe Liu, Zhicheng Liu, Junyan Yang, Qiao Wang.

Performed the mathematical modelling: Qinghe Liu, Zhicheng Liu, Qiao Wang. Analyzed the data: Qinghe Liu, Deqiang Li, Zhicheng Liu.

Collect the data: Zefei Gao, Junkai Zhu. Wrote the paper: Qinghe Liu, Zhicheng Liu.

All authors read and approved the final manuscript.

## Acknowledgments

Not applicable.

## Appendix1 Data Source (All data used in paper is public.)

### Reported Cases Data

https://www.who.int/

https://www.cdc.gov/

http://www.nhc.gov.cn/

http://wjw.beijing.gov.cn/

http://wjw.fujian.gov.cn/

http://wsjk.tj.gov.cn/

http://wsjkw.hebei.gov.cn/

http://www.xjhfpc.gov.cn/

http://wsjkw.nx.gov.cn/

http://ynswsjkw.yn.gov.cn/wjwWebsite/web/index

http://www.gzhfpc.gov.cn/

http://wsjkw.sc.gov.cn/

http://wsjkw.cq.gov.cn/

http://wst.hainan.gov.cn/swjw/index.html

http://wsjkw.gxzf.gov.cn/

http://wsjkw.gd.gov.cn/

http://wjw.hunan.gov.cn/

http://wjw.hubei.gov.cn/

http://www.hnwsjsw.gov.cn/

http://wsjkw.shandong.gov.cn/

http://hc.jiangxi.gov.cn/

http://wjw.ah.gov.cn/

http://www.zjwjw.gov.cn/

http://wjw.jiangsu.gov.cn/

http://wsjkw.sh.gov.cn/

http://wsjkw.jl.gov.cn/

http://wjw.nmg.gov.cn/

http://wjw.beijing.gov.cn/

http://sxwjw.shaanxi.gov.cn/

http://wjw.shanxi.gov.cn/

http://wsjk.ln.gov.cn/

http://wsjk.gansu.gov.cn/

http://wsjkw.hlj.gov.cn/

https://wsjkw.qinghai.gov.cn/

http://wjw.xizang.gov.cn/

https://weibo.com/

### Provincial-Level Migration Data

http://qianxi.baidu.com/

